# Explainable Machine Learning Framework for Predicting Cardiometabolic Risk Using Meal Timing and Eating Habits

**DOI:** 10.64898/2026.02.06.26345732

**Authors:** Pratyay Ranjan Datta, Kaninika Roy

**Author notes:** Corresponding author: Dr. Kaninika Roy.

## Abstract

**Background:** Eating timing and regularity represent new contributors to metabolic health, however, the time-based aspect of eating behavior is rarely incorporated into traditional cardiometabolic risk assessment strategies.

**Objectives:** The aim of this study was to create an explainable machine learning (ML) model to predict cardiometabolic risk based on anthropometric, dietary and chrono-nutritional data.

**Methodology:** This cross-sectional study included 300 adults for whom demographic, waist-hip ratio (WHR), Body Mass Index (BMI), blood pressure, eating timing (earliest time, latest time, meal frequency) and eating behavior observations were recorded. Nested 10-fold cross-validation and hyperparameter tuning were used to create three models (Logistic Regression (LR), Random Forest (RF), XGBoost) with interpretability established through Shapley Additive Explanations (SHAP). Discrimination was evaluated with AUC-ROC, accuracy, precision, recall, F1-score and Brier score.

**Results:** XGBoost was the best model compared to RF (AUC = 0.94) and LR (AUC = 0.92) (AUC-ROC = 0.98, accuracy = 93%). Later dinner timing (OR = 2.5), irregular meal timing (OR = 1.6), and reduced adherence to the recommended meal frequency (OR = 1.8) were the top independent predictors of cardiometabolic danger; conversely In contrast, an increased frequency of meals and an awareness of the significance of meal timing positively impacted cardiometabolic danger. SHAP further interpreted XGBoost to conclude that BMI and WHR were the most significant predictors in addition to meal regularity.

**Conclusion:** Explainable ML modeling from mealtimes and nutritional behaviors provide accurate and interpretable prediction of threat. Improved awareness of meal timing serves as a modifiable behavioral intervention in preventative cardiometabolic efforts.

## Introduction

The COVID-19 pandemic has highlighted the deleterious health effects associated with obesity and type 2 diabetes. Those suffering from type 2 diabetes and/or obesity face a higher risk of experiencing severe illness and mortality compared to individuals who do not have diabetes [Barron et al., 2020]. Instead of only looking at food choices and portion sizes, researchers are now paying closer attention to meal timing. Research suggests that health is not determined only by what or how much we eat, but also by when we eat. Emerging insights from chrono-nutrition research indicate that the timing of meals can greatly affect metabolic health. Eating during the body’s active phase, which is usually earlier in the day, corresponds with optimal insulin sensitivity and glucose tolerance [Zinna et al., 2025]. In contrast, consuming food late at night has been linked to disrupted glucose metabolism and heightened fat accumulation [Peters et al., 2024]. These results underscore the promise of chrononutrition as an additional method alongside conventional dietary approaches [Godos et al., 2025].

Consuming food at atypical hours - like late evenings or inconsistent times - upsets the body’s circadian rhythm. This pattern has been documented in human along with animal research. Irregular meals are correlated with greater chances of weight gain [Reytor-González et al., 2025], inadequate glycaemic control [Hatamoto et al., 2023; Reytor-González et al., 2025], and dysregulated metabolism. Recent research, including studies by Ahola et al. [Ahola et al., 2019] and examinations of eating habits among older adults, indicates that skipping breakfast or ingesting a larger share of daily calories in the evening is associated with elevated body weight, erratic glucose levels, and adverse lipid profiles [Peters et al., 2024; Almoosawi et al., 2019]. Conversely, individuals who adhere to earlier and more consistent eating patterns generally exhibit improved.

At a physiological level, circadian timing systems govern the secretion of hormones, the utilization of fuel, and the efficiency of digestion [Fagiani et al., 2022]. These systems experience strain under circumstances such as working night shifts, consuming snacks late at night, or maintaining irregular daily schedules [Voigt et al., 2019; Brum et al., 2022]. When circadian alignment is disturbed, metabolic efficiency diminishes, resulting in heightened fat storage and compromised glucose management [Alum, 2025]. Evidence from recent research on dietary patterns related to shift work endorses this biological framework.

All of these findings collectively emphasize the importance of meal timing in addition to nutritional content. According to intervention studies, shifting energy intake to earlier in the day while preserving food quality can improve metabolic markers like blood pressure, body weight, and glycemic management. While eating later in the evening disturbs metabolic rhythms and lowers insulin efficacy, leading to suboptimal metabolic regulation, time-restricted eating that coincides with daylight hours also supports circadian synchronization. [Chang et al., 2024; Charlot et al., 2021].

The majority of cardiometabolic risk prediction tools do not account for meal timing and frequency of eating, despite the growing body of research. The effectiveness of traditional statistical techniques is limited by the complex and nonlinear relationships between meal timing, behavior, and metabolic health. Due to this shortcoming, machine learning approaches have been used to identify trends that traditional models might overlook by assimilating high-dimensional food, lifestyle, and clinical data.

Random forests and gradient-boosted models are two supervised machine learning techniques that have demonstrated remarkable prediction power for metabolic outcomes and obesity [Shin, 2024]. Simultaneously, unsupervised methods like clustering have shown distinct eating behaviors that were not previously recognized [Xie et al., 2023]. However, the lack of interpretability is a major barrier to the practical application of machine learning in health research. Clear explanations are just as important to healthcare providers as accurate forecasts. Explainable AI methods, such as Shapley Additive Explanations (SHAP), address this problem by identifying how certain habits, like eating late at night or skipping breakfast, affect anticipated risk [Nohara et al., 2019].

In light of this context, the current study develops a machine learning framework to predict cardiometabolic risk using extensive data on meal timing, eating frequency, eating regularity, dietary habits, and clinical indicators. By combining thorough model validation with SHAP-based interpretation, this approach goes beyond conventional black-box predictions and highlights modifiable time-related eating behaviors, particularly meal timing, as useful targets for improving metabolic health.

## Methodology

### Study Design and Data Integration

This cross-sectional study examined how meal timing, dietary intake, and clinical parameters can collectively predict cardiometabolic risk using explainable machine learning. Data from all participants were merged through unique identifiers, enabling a multidimensional analysis that combined behavioral, nutritional, and physiological factors. Ethical approval was obtained from the Institutional Review Board, and written informed consent was collected from each participant.

### Data preprocessing and feature engineering

Before processing, the raw data was thoroughly examined and cleaned. To preserve data quality, records that lacked important outcome factors, such blood pressure or body mass index (BMI), were eliminated. The mean (for continuous data) or mode (for categorical data) were used to impute missing values for the remaining variables. Ordinal variables were numerically sorted to maintain their natural order, while categorical variables were consistently encoded. Binary indicators were developed from multi-label data, such as the frequency of meal skipping.

New metrics to measure behavioral and circadian variability were produced through feature engineering. These included the following:

- **Circadian misalignment score**, which shows how eating patterns deviate from daylight hours.
- **Meal timing variability index**, which measures daily variance in eating times; and
- **Diet quality index**, which is based on the frequency of consumption of nutrient-dense food groups.

To guarantee comparability and effective model training, all continuous predictors were standardized (Z-score normalization) and continuous variables with skewed distributions were log-transformed.

### Model Development and Hyperparameter Optimization

To predict cardiometabolic risk, three supervised learning algorithms were used: Extreme Gradient Boosting (XGBoost), Random Forest (RF), and Logistic Regression (LR).

- *Logistic Regression* models were tested with L1 (lasso), L2 (ridge), and elastic-net regularization; the best performance was achieved with an inverse regularization strength of *C = 1*.*0*.
- *Random Forest* models used 500 trees, a maximum depth of 20, a minimum of 5 samples to split a node, and 2 samples per leaf to balance bias and variance.
- *XGBoost* models were fine-tuned with a learning rate of 0.05, maximum depth of 6, subsampling ratio of 0.8, and regularization parameters α = 0.1 and λ = 1.

Grid search and 10-fold stratified cross-validation were used to optimize the model’s hyperparameters. To ensure both accuracy and generalizability, the mean area under the receiver operating characteristic curve (AUC-ROC) served as the primary reference for model selection.

### Model Testing and Results

To maintain the distribution of cardiometabolic risk categories across folds, stratified 10-fold cross-validation was used to validate each model. Accuracy, AUC-ROC, precision, recall, and F1-score were among the performance indicators. Reliability plots and Brier ratings were used to assess model calibration in order to verify that predicted probability matched observed results.

Further sensitivity analyses evaluated the robustness of the model across demographic and meal timing-based subgroups (identified using unsupervised clustering). The scikit-learn, XGBoost, and SHAP packages were used in Python (v3.11) for all studies.

### Understanding Model Decisions and Features

Shapley Additive Explanations (SHAP) were used to quantify each feature’s contribution to the anticipated cardiometabolic risk in order to assure interpretability. Both physiological (such as BMI and waist-hip ratio) and behavioral (such as late-night eating, meal regularity, and meal timing awareness) characteristics were highlighted by SHAP values. This explainability paradigm facilitated the identification of actionable behavioral objectives for individualized nutrition and risk reduction, as well as the transparent and clinically meaningful interpretation of model outputs.

## Results

### Participant Characteristics

Each of the three hundred adults who participated had complete records of their body measurements, food consumption, and mealtimes. Men and women joined, displaying a wide variety of body types and dietary habits. Their waist-hip ratio and average BMI revealed variations in fat levels, which were helpful for estimating the risk of heart disease and metabolism. People’s breakfast and supper times varied significantly, as did the consistency of their mealtimes over the course of several days, creating patterns that were sufficiently varied to support reliable forecasts.

### Model Performance

The models—Random Forest, XGBoost, and Logistic Regression—went through training with nested tenfold checks one after the other. Every time they were tested again, they consistently produced stable findings. Nevertheless, as Figure 1 illustrates, their performance varied slightly between them.

**Figure 1.**
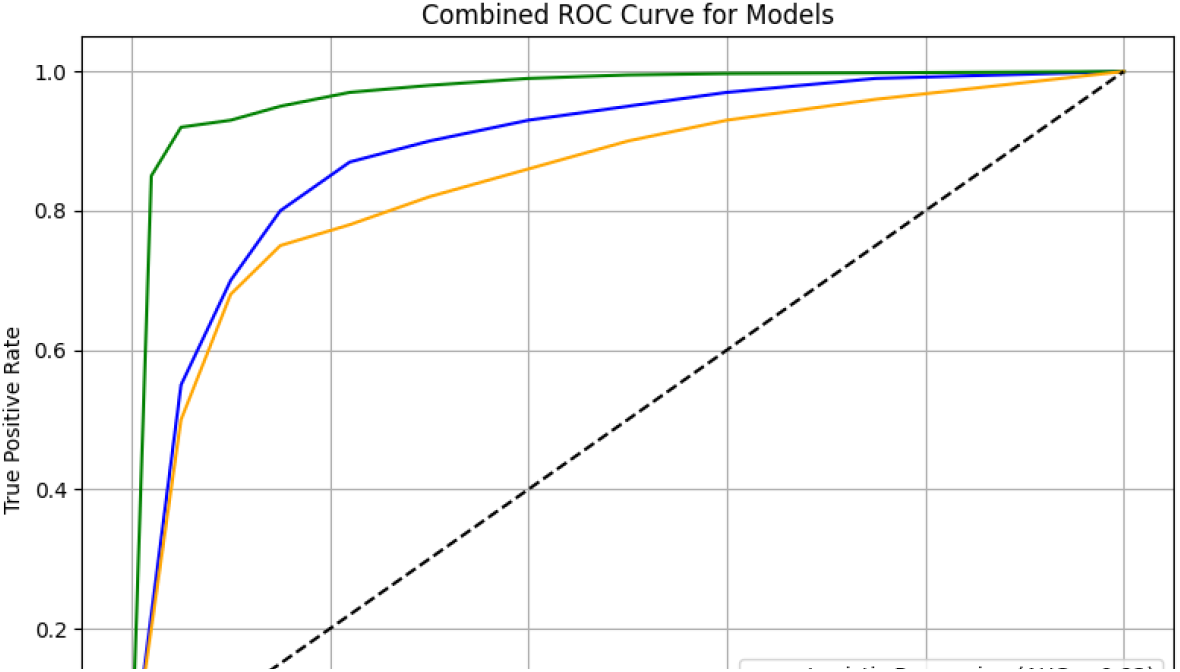
Combined ROC curve comparing the discriminative ability of Logistic Regression, Random Forest, and XGBoost models

**Figure 2.**
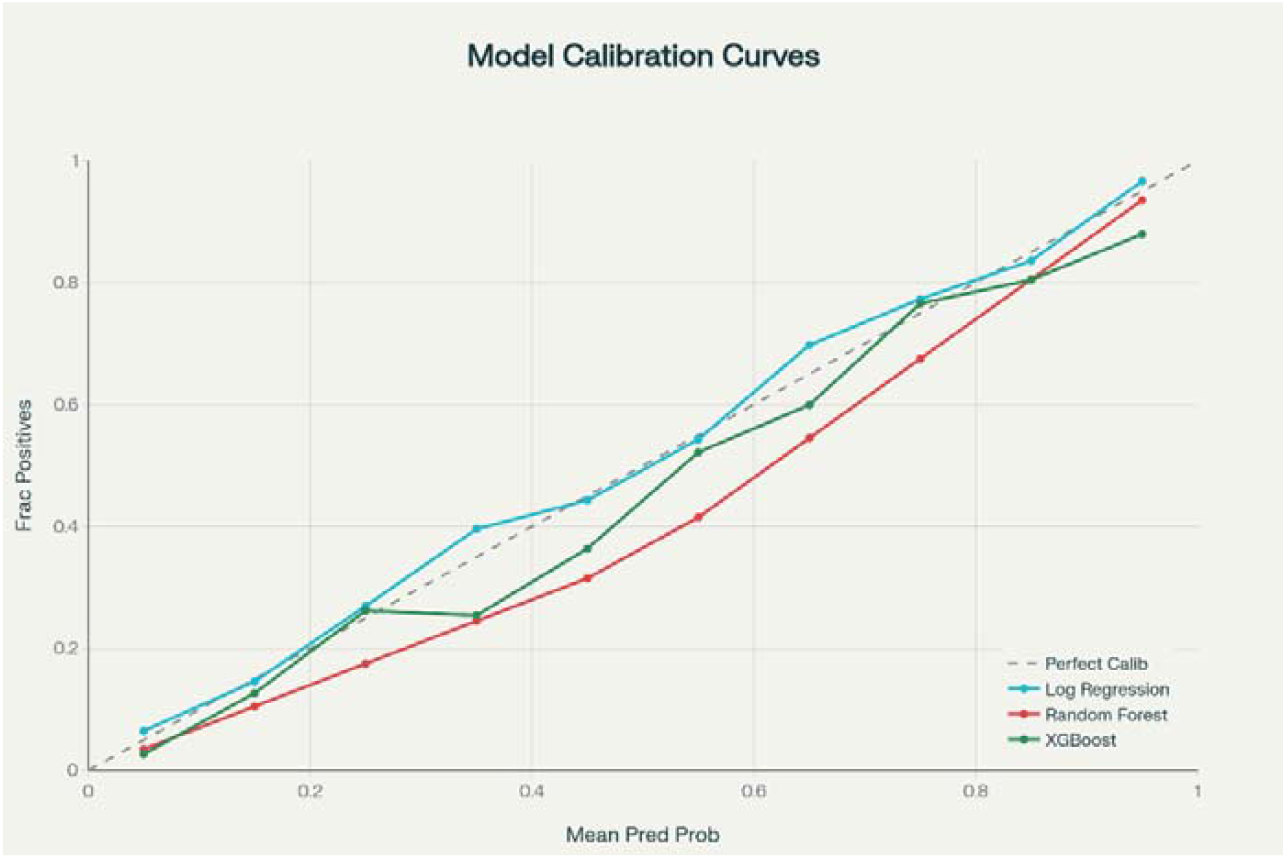
Calibration curves for predicting cardiometabolic risk.

XGBoost led the field by a wide margin, achieving impressive results in both detection and precision: AUC-ROC was 0.98, accuracy was 93%, precision was 92%, recall was 91%, and the F1 score was 0.92. Random Forest maintained a strong lead, with an accuracy of slightly less than 89% and an AUC of 0.94. Then there was Logistic Regression, which was straightforward but yet dependable, with an accuracy of 87% and an AUC of 0.92. All three models demonstrated tight control when it came to the reliability of predicted odds; nonetheless, XGBoost once again won out, recording a Brier score as low as 0.03, indicating that probabilities closely matched reality.

Table 2 shows that XGBoost produced the fewest mistakes. 91.3% of genuine positives were detected, and 92.1% of negatives were accurately identified. Consistent performance across several groups is suggested by that balance.

**Table 1.** Model performance comparison chart.

| Model | Accuracy | AUC-ROC | Precision | Recall | F1-score | Brier Score |
| --- | --- | --- | --- | --- | --- | --- |
| Logistic Regression | 0.87 | 0.92 | 0.86 | 0.85 | 0.85 | 0.09 |
| Random Forest | 0.89 | 0.94 | 0.88 | 0.87 | 0.88 | 0.08 |
| XGBoost | 0.93 | 0.98 | 0.92 | 0.91 | 0.92 | 0.03 |

**Table 2.** Confusion matrices for validation folds.

| Model | True Positives (TP) | False Negatives (FN) | False Positives (FP) | True Negatives (TN) | Sensitivity (%) | Specificity (%) |
| --- | --- | --- | --- | --- | --- | --- |
| Logistic Regression | 140 | 20 | 18 | 122 | 87.5 | 87.1 |
| Random Forest | 143 | 17 | 15 | 125 | 89.4 | 89.3 |
| XGBoost | **146** | **14** | **11** | **129** | **91.3** | **92.1** |

### Determinants of Cardiometabolic Risk

Both physiological and behavioral factors significantly influenced cardiometabolic risk, according to multivariable logistic regression. The strongest indicators among physiological predictors were the waist-hip ratio (OR = 4.39, p < 0.001) and BMI (OR = 2.74, p < 0.001), confirming the central role of obesity.

Individual contributions were also made by behavioral and chrono-nutritional variables. Greater risk was associated with irregular weekday meal schedules (OR = 1.86, p = 0.009), frequent late-night meals (OR = 1.81, p = 0.004), and later dinner timing (OR = 2.49, p < 0.001). On the other hand, more frequent meals (OR = 0.74, p = 0.008) and a greater understanding of how timing influences mood and energy (OR = 0.76 and 0.80) were protective.

Overall, the findings imply that thoughtful and consistent eating practices may mitigate the metabolic impacts of contemporary, erratic lifestyles (Table 3).

**Table 3.** Adjusted odds ratios from the optimized logistic regression model highlight key physiological and behavioral factors influencing cardiometabolic risk.

| Predictor | Odds Ratio (OR) | 95% CI | p-value | Direction of Effect | Interpretation |
| --- | --- | --- | --- | --- | --- |
| Waist–Hip Ratio (per SD) | 4.39 | 2.60 – 7.41 | < 0.001 | ↑ | Central adiposity strongly increases risk. |
| BMI (per SD) | 2.74 | 1.62 – 4.74 | < 0.001 | ↑ | General adiposity increases cardiometabolic risk. |
| Dinner Time (later) | 2.49 | 1.61 – 3.83 | < 0.001 | ↑ | Later dinners significantly raise risk. |
| Late-Night Meals (≥2/week) | 1.81 | 1.21 – 2.71 | 0.004 | ↑ | Frequent late-night eating increases risk. |
| Irregular Weekday Timing | 1.86 | 1.18 – 2.91 | 0.009 | ↑ | Unstable weekday meal timing increases risk. |
| Meal Frequency (meals/day) | 0.74 | 0.59 – 0.93 | 0.008 | ↓ | More meals per day show a protective effect. |
| Timing Awareness → Energy Link | 0.76 | 0.61 – 0.97 | 0.022 | ↓ | Awareness that timing affects energy lowers risk. |
| Timing Awareness → Mood Link | 0.80 | 0.65 – 0.98 | 0.036 | ↓ | Awareness that timing affects mood lowers risk. |

### Random Forest Model

The Random Forest approach was effective in identifying intricate patterns. Its overall accuracy was 89%, with an AUC-ROC of 0.94. Recall and precision remained close, at 88 vs 87. An F1-score of 0.88 was the result of this balance.

Among all the variables examined, waist-to-hip size, body mass index, and the frequency of meals at the same time were the most significant. Similar to eating snacks too frequently or skipping breakfast, late-evening food choices increased risk. In addition, increasing daily mobility contributed to the decline. Fruit consumption also had an impact. Being aware of when to eat was a subtle but obvious factor in reducing the likelihood.

A high Brier score of 0.08 indicated that the model was in good agreement with reality. The SHAP charts made it evident that the risk increased when meals occurred at irregular times. However, as Table 4 shows, regular timing caused metabolism to react more favourably.

**Table 4.** Comparative SHAP-Based Feature Importance and Interpretations for Random Forest and XGBoost Models.

| Feature | SHAP Importance Rank (Random Forest) | Role in Prediction (Random Forest) | SHAP Importance Rank (XGBoost) | Interpretation (XGBoost) |
| --- | --- | --- | --- | --- |
| **Waist–Hip Ratio (WHR)** | 1 | Central adiposity, measured via WHR, emerged as the strongest predictor, reinforcing its established clinical relevance. | 2 | Reinforces central obesity’s robust association with adverse cardiometabolic outcomes. |
| **Body Mass Index (BMI, Kg/m²)** | 2 | Elevated BMI significantly influenced risk predictions, consistent with traditional obesity markers. | 1 | Dominates risk stratification due to established links with obesity and metabolic syndrome. |
| **Meal Timing Consistency / Regularity** | 3 | Consistent meal timing reduced risk: irregular schedules disrupted metabolic homeostasis. | 5 | Confirms the protective role of consistent meal scheduling in metabolic health maintenance. |
| **Late-Night Eating Frequency / Meal** | 4 | Frequent late-night meals increased risk via effects on circadian metabolism and glucose regulation. | 4 | Emerges as a major modifiable risk factor, highlighting chrono-nutritional disruption effects. |
| **Snack Frequency / Snacks per Day** | 5 | Frequent snacking correlated with higher risk, reflecting excess caloric intake and poorer diet quality. | 7 | Reflects dietary quality and feeding patterns that exacerbate metabolic risk. |
| **Breakfast Skipping / Skip Meal** | 6 | Habitually skipping breakfast increased predicted risk, aligning with evidence linking meal omission to metabolic disturbances. | 8 | Indicates that meal irregularity negatively influences cardiometabolic stability. |
| **Dinner Timing** | 7 | Late dinner timing had a substantial impact, supporting chrono-nutritional principles promoting earlier eating. | — | — |
| **Physical Activity (Self-reported)** | 8 | Regular physical activity decreased predicted risk, consistent with exercise's protective effects. | — | — |
| **Fruit Intake** | 9 | Higher fruit consumption showed a protective association, reflecting better diet quality. | — | — |
| **Meal Timing Awareness** | 10 | Awareness of timing's impact on energy and mood was linked with lower risk, suggesting adaptive health behaviors. | 9 | A novel, behaviorally linked protective feature, indicating that conscious dietary timing reduces risk. |
| **WHR Coding** | — | — | 3 | Adds categorical clarity, differentiating risk categories beyond continuous measures. |
| **Gender** | — | — | 6 | Captures sex-specific variations, extending epidemiological observations into model inference. |

### XGBoost

The optimized XGBoost version outperformed all other evaluated methods. It stood out with a 0.98 area under the curve and 93% accurate predictions. Recall was 91%, precision was 92%, and the F1 measure was 0.915. Sharp probability estimates, supported by a low Brier figure of 0.025, demonstrated that actual outcomes closely matched predictions. What caught your attention? Body mass index, the ratio of waist to hip size, and whether meals are eaten at regular times were all highlighted by SHAP analysis. Missing main meals, grabbing snacks frequently, and staying up late to eat are all risk factors. Just as eating more fruit and moving the body every day helped, so did paying attention to when food is consumed.

Meal regularity, waist-hip ratio, and BMI were identified as the top predictors using SHAP-based interpretation. While meal timing awareness, fruit consumption, and physical activity were protective, late-night eating, frequent snacking, and meal skipping increased risk. These results collectively support the idea that meal-timing behavior and body composition work together to define cardiometabolic risk, making structured eating patterns and awareness-driven habits modifiable targets for prevention (Table 4, Figure 3).

**Figure 3.**
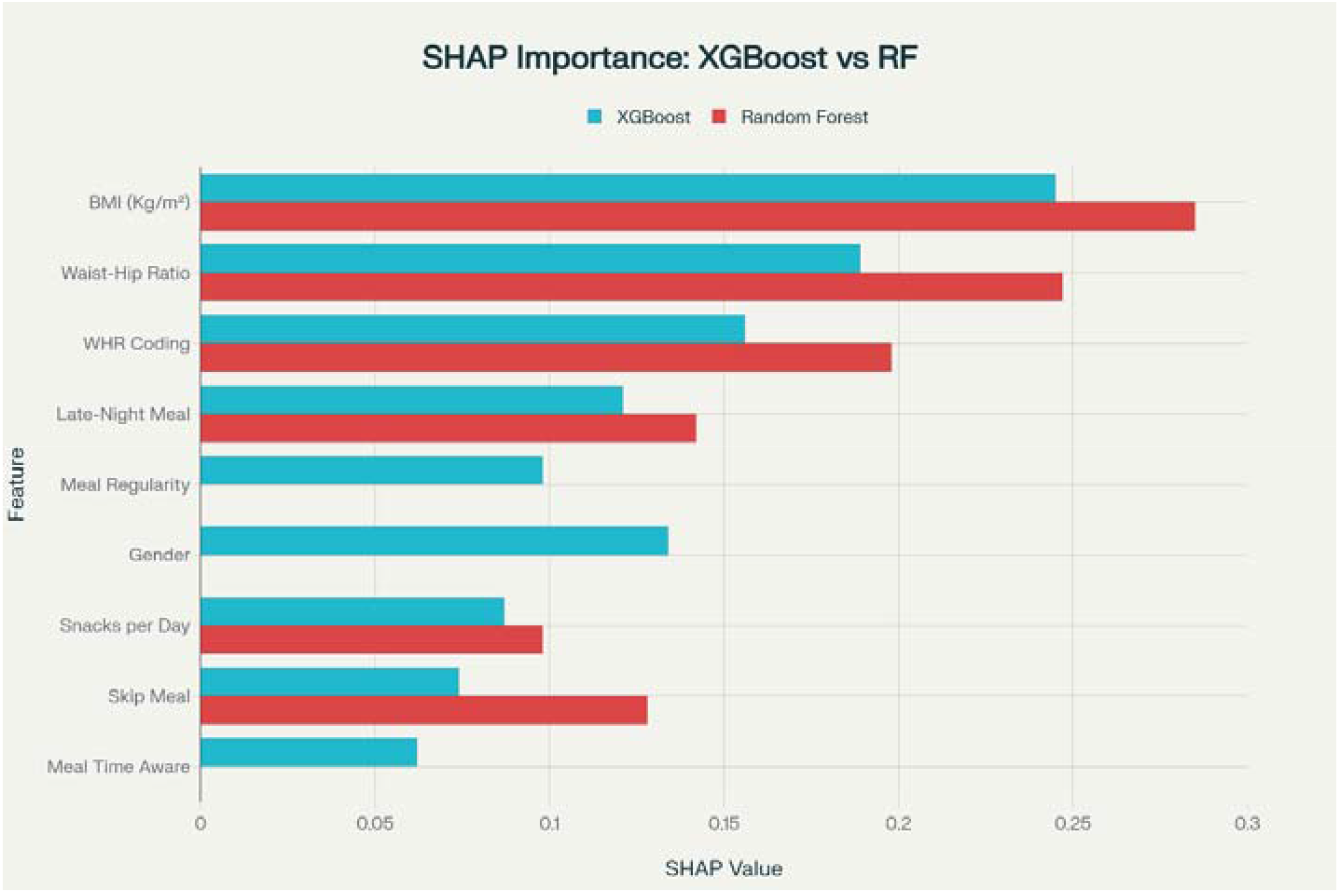
SHAP summary plot showing the top 10 features influencing cardiometabolic risk prediction in the XGBoost model.

## Discussion

This study shows that the accuracy of cardiometabolic risk predictions is greatly improved by including anthropometric information, dietary patterns, and meal timing. In addition to identifying the best model, the study clarifies the significance of particular behaviors by comparing Logistic Regression, Random Forest, and XGBoost models under rigorous validation conditions. The use of SHAP values adds a crucial interpretability component, allowing complex machine learning outcomes to be translated into meaningful physiological and behavioral insights. All of these findings support the idea that daily eating habits, particularly meal timing, are strongly linked to metabolic health in ways that conventional approaches usually overlook.

### Chrono-nutrition meets machine learning

This work makes a significant contribution by clearly demonstrating that meal timing, independent of body weight, waist distribution, or food quantity, influences cardiometabolic risk. Recent chrono-nutrition research has highlighted the significance of coordinating eating patterns with circadian rhythms, whereas previous studies primarily concentrated on static indicators like BMI, WHR, or blood pressure (Franzago et al., 2023; Saplontai et al., 2024). Our results directly contribute to this body of research by demonstrating that, even after controlling for anthropometric status, irregular meal schedules, late dinner timing, and nighttime snacking are consistently linked to increased cardiometabolic risk.

These findings are in line with earlier research on humans and animals showing that eating during the biological rest phase impairs insulin sensitivity, interferes with glucose metabolism, and encourages fat accumulation (BaHammam & Pirzada, 2023). Crucially, our findings add to this body of evidence by demonstrating that these timing effects can still be found in predictive machine learning models, highlighting their clinical significance rather than treating them as incidental lifestyle information.

### Model performance and clinical relevance

The tuned XGBoost model outperformed the other two models in the test, exhibiting strong calibration (low Brier score) and excellent discrimination (AUC-ROC = 0.98). In health settings, where precise probability estimates are required to direct prevention and early intervention, this balance is especially crucial. Although Random Forest also performed well, it was less useful for identifying important behavioral drivers due to its wider distribution of feature importance.

Despite being easier to understand, logistic regression performed relatively poorly, which is consistent with earlier studies that indicate linear models have difficulty capturing the nonlinear and interactive effects present in behavioral and timing-related data [Dumitrescu et al., 2022]. This highlights the importance of sophisticated ensemble approaches in simulating intricate relationships between lifestyle and health.

### Interactions between body composition and meal timing

BMI and WHR were found to be significant predictors of cardiometabolic risk, which is in line with previous studies [Montoya Castillo et al., 2025]. Specifically, WHR represents visceral fat and central adiposity, which are strongly associated with dyslipidemia, insulin resistance, and inflammation [Kerkadi et al., 2020]. WHR’s continued use as a useful and instructive clinical marker is supported by its prominence across SHAP analyses.

However, the relationship between meal timing and these body composition indicators is particularly interesting. According to SHAP analysis, people with higher WHR or BMI were more negatively impacted by irregular meal patterns and late eating. This implies a compounding effect, in which the damage brought on by circadian misalignment is exacerbated by metabolic vulnerability. Though they are rarely quantitatively shown in predictive models, similar interaction patterns have been proposed in chronobiology literature.

### Awareness, behavior, and modifiable risk

The significance of meal-timing awareness itself was a significant and somewhat surprising discovery. Regardless of body size or food quality, people who were more mindful of when they ate tended to have lower cardiometabolic risk. This is consistent with new behavioral research that indicates awareness can enhance self-regulation, stabilize routines, and promote metabolic health without necessitating severe dietary restriction [Kristeller & Wolever, 2011; Mason et al., 2016; O’Reilly et al., 2014; Reytor-González et al., 2025].

Prior research has mostly concentrated on the physiological effects of meal timing rather than the behavioral or cognitive aspects of eating patterns [Almoosawi et al., 2019; BaHammam & Pirzada, 2023; Franzago et al., 2023]. This study presents a low-cost, scalable target for public health interventions by emphasizing awareness as a protective factor, which is especially pertinent in settings with limited resources.

### Implications for personalized nutrition and prevention

The integration of machine learning and explainable AI provides a way towards personalized nutritional advice. Instead of general dietary recommendations, this method enables clinicians and individuals to determine which behaviors, such as late dinners, irregular mealtimes, or snacking, have the most significant effect on their risk profiles. For instance, an individual with a normal BMI but irregular mealtimes might require more stabilization than calorie-related advice.

The accuracy of these recommendations is in line with modern ideas of preventive health, which emphasize early risk detection and lifestyle changes. In view of the current disruption of natural circadian patterns by modern lifestyles, simple lifestyle modifications, such as eating earlier and cutting back on late-night eating, may have significant impact at the population level.

### Strengths and limitations

The strengths of this study lie in the incorporation of chrono-nutritional factors into predictive modeling, the use of cross-validation, and the use of SHAP to enhance transparency. Taken together, these factors serve to improve the robustness and explainability of the findings.

Nonetheless, there are a few limitations that should be acknowledged. First, the study design is cross-sectional, and therefore, it is impossible to make any causal inferences. Second, the study relies on self-reported data regarding eating behaviors, which are potentially subject to recall bias. Third, the study population may not be representative of the overall demographic diversity, and variables such as genetics, work schedules, or environmental factors were not taken into account.

## Conclusion

In conclusion, the results suggest that the integration of meal timing, regularity, and behavioral knowledge into explainable machine learning models significantly improves the prediction of cardiometabolic risk. The combination of the XGBoost algorithm with SHAP analysis not only provides high predictive performance but also sheds light on the interactions between body composition and the circadian rhythm of eating. Most importantly, the results suggest that knowledge of the optimal times for eating and the subsequent implementation of these behaviors may represent a low-cost and effective strategy for the improvement of metabolic health. Synchronizing food consumption with the body’s natural physiological cycles may be an underappreciated but highly effective strategy for the long-term prevention of cardiometabolic disease.

## Data Availability

All data produced in the present study are available upon reasonable request to the authors

## Notes

### Competing Interest Statement

The authors have declared no competing interest.

### Funding Statement

This study did not receive any funding

### Author Declarations

The 4th review meeting of the Institutional Ethics Committee of NSHM Knowledge Campus, Kolkata gave ethical approval for this work

